# Using Polygenic Scores for Circadian Rhythm to predict Wellbeing, Depressive Symptoms, Chronotype, and Health

**DOI:** 10.1101/2023.06.02.23290377

**Authors:** A. Landvreugd, R. Pool, M. Nivard, M. Bartels

**Affiliations:** Department of Biological Psychology, Vrije Universiteit Amsterdam, The Netherlands; Amsterdam Public Health Research Institute, Amsterdam University Medical Centres, Amsterdam, The Netherlands

**Keywords:** Circadian Rhythm, Polygenic Scores, Twins, Mental Health, Wellbeing, Depression

## Abstract

The association between the circadian rhythm and diseases has been well-established, while the association with mental health is less explored. Given the heritable nature of the circadian rhythm, this study aimed to investigate the relationship between genes underlying the circadian rhythm and mental health outcomes, as well as a possible gene-environment correlation for circadian rhythm. In a sample from the Netherlands Twin Register (*N* = 14,021), polygenic scores (PGSs) were calculated for two circadian rhythm measures: Morningess and Relative Amplitude. The PGSs were used to predict mental health outcomes such as subjective happiness, quality of life, and depressive symptoms In addition, we performed the same prediction analysis in a within-family design in a subset of dizygotic twins. The PGS for Morningness significantly predicted Morningness (*R*^2^ = 1.55%,) and Depressive Symptoms (*R*^2^= 0.22%,). The PGS for Relative Amplitude significantly predicted General Health (*R*^2^ = 0.12%,) and Depressive Symptoms (*R*^2^ = 0.20%,). Item analysis of the depressive symptoms showed that 4/14 items were significantly associated with the PGSs. The within-family results hinted at a gene-environment correlation for Morningness. Overall, the results showed that people with a genetic predisposition of being a morning person or a high relative amplitude are likely to have fewer depressive symptoms. Contrarily to our hypotheses, the four associated depressive symptoms described symptoms related to decision-making, energy, and feeling worthless, rather than sleep. Our findings plead for a substantial role for the circadian rhythm in depression research, and to further explore the gene-environment correlation in the circadian rhythm.

## Introduction

Since the introduction of electric lighting in the early 19^th^ century, human exposure to light has changed drastically (Falchi et al, 2016). Artificial lighting has enabled us to be exposed to light whenever we want, but also when we do not. Streets are lit after sunset and our phones and laptops only stop emitting light when we decide to turn them off. It is plausible, perhaps even likely, that these changes affect our circadian rhythm, the body’s ‘biological clock’. The circadian rhythm is a biological process that runs in proximately twenty-four hours cycles to carry out essential functions, such as regulating sleep, temperature, and digestion (Reddy et al., 2021). The circadian rhythm is thought of as endogenous, but is synchronized by several environmental factors, so-called ‘zeitgebers’, of which the most influential one is light exposure (Murray et al., 2020).

Due to the increasing popularity of electric lighting over the centuries, the functioning of the circadian rhythm may have been disrupted (Wright et al., 2001, 2013). To measure the effects of a disrupted circadian rhythm, researchers use various circadian rhythm derivatives. Common derivates include chronotype, core body temperature, melatonin levels, and rest-activity cycles (Haffen, 2009), either in humans or in animal models. For example, studies in humans have found associations between melatonin levels and metabolic disorders (McHill et al., 2014; Shimada et al., 2016), such as insulin resistance (Tuomi et al., 2016). Other studies, also in humans, used sleep disorders as a measure and found links to cancer types (Hu et al., 2013; Papagiannakopoulos et al., 2016) or looked at sleep disturbances and found links to infertility (Baumgartner et al., 1993; Touzet et al., 2002). Studies in mice, for example, focused on a mutation in a particular gene associated with the circadian rhythm (e.g. Per2, Per3, Cry1, Ko & Takahashi, 2006). Viswambharan et al., (2007) and Wang et al., (2008) found associations for such genetic mutations with cardiovascular diseases (Per2). Finally, another common way to study the effects of a disrupted circadian rhythm is by studying shift workers. A meta-analysis analyzed twenty-one observational studies to find that night shift workers are more likely to suffer from type-2 diabetes (Gao et al., 2020a). Behrens et al., (2017) studied shift- and night-workers and found that they had a twofold hazard ratio of having prostate cancer.

Besides being linked to several physical conditions, circadian measures have also been associated with neurodegenerative diseases and mood disorders. For example, for a while the idea remained that circadian disruption was merely a symptom of Alzheimer’s disease and Parkinsons’s disease (McCurry et al., 1999; Shulman et al., 2001). Only recently studies have started to show that circadian disruption may work as a catalyst speeding up the progression of these diseases (Chen et al., 2015; Gu et al., 2015). Other studies have found that meaures of circadian disruption, specifically insomnia, are correlated to mood disorders such as depression and bipolar disorder (Lind et al., 2017; Murray et al., 2017).

In addition to environmental factors that influence the circadian rhythm, such as light, the role of genetic factors is becoming more evident. Twin studies have shown that chronotype has a heritability estimate between 40 – 54% (Barclay et al., 2014; Hur, 2007). Important to notice here is that the heritability of morningness changes with age. Specifically, the additive genetic influences on *finding it easy to be active in the morning rather than in the evening*, are 34% in middle adulthood and 44% in younger and older adults (Barclay et al., 2014). This decrease in genetic influences during middle adulthood is possible driven by increased work and family responsibilities with more dictated daily activity patters that suppress genetic predisposition. Besides heritability studies, there has also been work focused on specific genes related to the circadian rhythm (Ko & Takahashi, 2006). Besides those previously mentioned (Gu et al., 2015; Viswambharan et al., 2007; Wang et al., 2008), there are plenty more studies that investigate these specific genes, both in humans and in animal models (Ebisawa et al., 2001; Lou et al., 2018; Mishima et al., 2005).

The circadian rhythm is subjected to both genetic and environmental factors, but these factors do not act separately, they also depend on each other. This is called ‘gene-environment correlation’: your environmental exposure depends on your genetic makeup. Recently, Burns et al., (2022) considered this theory for their genome-wide-gene-by-environment study on daylight and chronotypes. They studied data from 280.987 participants, including data on their DNA, chronotype (self-report), and time spent outdoors (self-report). They reported a positive effect of daylight exposure on morningness, i.e. greater daylight exposure was associated with greater morningness. One genetic variant was found to moderate this effect, meaning that the positive effect of daylight exposure on morningness is enhanced in people that carry this genetic variant. Based on this finding, one of the author’s hypotheses is that people with a genetic predisposition to seek out more daylight also tend to be morning people.

The heritable influences on complex traits is driven by thousands of genetic variants with small effects, rather than a few genetic variants with large effects. With this in mind, Jones et al., (2019) performed a Genome-wide Association Meta-Analysis using data from 697,828 individuals from UK Biobank and 23andMe participants. They found 351 loci associated with chronotype and reported a single-nucleotide polymorphism heritability (h^2^snp) of 13.7%. The results from Genome-wide Association studies (GWAS) can also help us understand the associations between various traits. For example, a common practice is to use GWAS summary statistics to examine genetic correlations. Jones et al., (2019) reported genetic correlations between being a morning person and psychiatric traits. They found both positive genetic correlations (subjective wellbeing) and negative genetic correlations (e.g. schizophrenia, depressive symptoms and intelligence) with morningness, but they also found that BMI and Type 2 diabetes were not genetically correlated to morningness. As they ultimately were interested in establishing whether morningness had a causal relation with (mental) health outcomes, they applied mendelian randomization analyses to estimate causal effects. Their main findings suggested that a genetic predisposition for being a morning person may reduce the odds of developing schizophrenia and increase levels of subjective wellbeing. Using the same methods, Lane et al., (2016) suggested that increased genetic predisposition for ‘eveningness’, is correlated with and possibly causal for increased educational attainment (educational attainment increased by 7.5 per increase in ‘eveningness’ category).

Where GWAS helps us to identify genetic markers related to specific traits, another application of GWAS is to use these markers found in the analysis to calculate polygenic scores (PGSs). Not many studies have applied this approach to circadian rhythm data yet, but Fergusson et al., (2018) calculated PGSs for low relative amplitude. The PGSs were found to predict mood instability, major depressive disorder, and neuroticism, meaning that having a high genetic risk for low relative amplitude, reflecting a predisposition for a disrupted rest-activity rhythm, is associated with a higher risk of developing these psychiatric phenotypes. This was not the case for generalized anxiety disorder and bipolar disorder

We built on the work of Jones et al., (2019) and Ferguson et al., (2018) to further explore the genetic association between the circadian rhythm and mental health. To this end we constructed PGSs for two different measures of the circadian rhythm. In the current study the circadian measures used were chronotype (‘morningness’) and relative amplitude (RA). RA is the difference in activity between the most active 10-h period and the least active 5-h period in a complete 24-h period. This means that high RA indicates a bigger difference between activity levels during the most active and least active periods of the day, compared to low RA. Low RA is indicative of a disrupted rest-activity rhythm. A PGS for self-reported chronotype, a subjective measure of circadian rhythm, and a PGS for RA as an objective measure of circadian rhythm were constructed in a sample of the Netherlands Twin Register. These two types of PGS were used to predict several outcomes such as wellbeing, depressive symptoms, chronotype, and health.

To enable detection of possible gene-environment correlation in the associations between circadian rhythm and sleep, wellbeing and mood related outcomes, the analyses were extended from a between-family design to a within-family design that only included dizygotic twins. The theory behind this method is that the twins all grew up in the same family environment, so you expect the effect of the family environment to be zero in that sample, being left with only genetic effects. In the between-family sample, you expect to have both genetic and family effects. If the prediction results from both samples are significantly different from each other, one can conclude that there is gene-environment correlation at play.

## Methods

### Participants

Participants in this study voluntarily registered to participate in scientific research at the Netherlands Twin Register (NTR, Ligthart et al., 2019). The NTR sample is a population-based sample of twins and their families. On average every two to three years a survey is send out designed to measure, among other things, personality, psychopathology, wellbeing, and lifestyle.

In this study, we used data from 14021 adult participants (mean age 43.01, *SD* = 17.63, 63.3% female) for whom phenotype and genotype data were available. The data came from surveys (‘waves’) 2 (1993-1994), 3 (1995-1996), 4 (1997-1998), 8 (2009-2010), 10 (2013-2014), and 14 (2019-2020). In the event of multiple available time-points per individual for the same measure, the last time-point was selected.

### Outcome measures

#### Wellbeing

*Hedonic wellbeing* is assessed with the Subjective Happiness Scale (Lyubomirsky and Lepper., 1999), the Satisfaction with life scale (Diener et al., 1985), and the Cantril ladder (Cantril., 1965). The Subjective Happiness Scale is a four-item survey where the items are rated on a Likert scale from 1 (strongly disagree) to 7 (strongly agree). An example item is: ‘Compared with most of my peers, I consider myself more happy’. We recoded two negatively worded items, so that for all items a higher score was associated with a higher level of happiness. The Satisfaction with Life Scale (Diener et al. 1985) is a five-item survey where the items are also rated on a Likert scale from 1 (strongly disagree) to 7 (strongly agree). An example item is: ‘*So far I have gotten the important things I want in life’*. The Cantril Ladder (Cantril, 1965) invites participants to rate their quality of life on a 10-point scale, where score of 0 represents the worst life possible, whereas a score of 10 represents the best life possible (‘Where on the scale would you put your life in general?’)

*Eudaimonic wellbeing* is assessed with the Short Flourishing Scale (Diener et al., 2010), which consists of 8 items that participants rated from 1-7 (‘strongly disagree’ – ‘strongly agree’) on a Likert scale. An example item is: ‘I lead a purposeful and meaningful life’.

#### Health

*Self-rated health* was assessed using a single item: ‘How would you rate your general health?’ (Eriksson et al., 2001,). Participants rated the item on a 5-point Likert scale ranging from ‘Bad’ to ‘Excellent’.

#### Chronotype

A single item was used to assess *Chronotype*: ‘Are you a morning-active or evening-active person?’ The scale ranges from 1-5 with 1 representing a morning-active person and 5 representing an evening-active person.

### Depressive symptoms

The adult self-report (ASR) of The Achenbach System of Empirically Based Assessment (ASEBA, Rescorla & Achenbach, 2004) was used to measure the DSM *depressive problems scale* (Achenbach & Rescorla, 2003). Fourteen items (see Table S1) were rated from 0-2 (0 = not true, 1 = somewhat true, 2 = very true). An example item is: ‘I cry a lot’. The data included the sum scores of the scale, as well as the scores on the individual items.

### GWAS summary statistics

To create the polygenic scores, GWAS summary statistics for two traits were used: morningness and relative amplitude.

### Morningness

We include morningness as a subjective measure of circadian rhythm and we used the publicly available summary statistics from the GWAMA by Jones et al. (2019). Using a four-point Likert scale, participants reported whether they identified as a morning person or an evening person. For this study, we used the summary statistics that only included UK Biobank participants (*N* = 449,734). We retained variants for which the effect allele frequency (EAF) was 0.01 ≤ EAF ≤ 0.99. Variant EAF and effect sizes were aligned with the Netherlands Twin Register (NTR) reference for the 1000 genomes variants. Discovery variants that are not part of this reference were discarded.

### Relative Amplitude (RA)

The publicly available summary statistics from the GWAS by Ferguson et al. (2018) were used to make PGSs for RA. RA was derived from accelerometer data and is therefore commonly used as an objective measure of circadian rhythm. The GWAS was performed in a sample of 71,500 UK Biobank participants. We retained variants for which the effect allele frequency (EAF) is 0.01 ≤ EAF ≤ 0.99. Variant EAF and effect sizes were aligned with the NTR reference for the 1000 genomes variants. Discovery variants that are not part of this reference were discarded.

### Statistical analyses

#### Polygenic score computation

The summary statistics from Jones et al. (2019) and Ferguson et al. (2018) were taken as input for the LDpred 0.9 software Vilhjálmsson et al., 2015). For estimating the target LD structure, we (1) used a selection of unrelated individuals in the NTR sample and (2) selected a set of well-imputed variants in the NTR sample. The parameter ld_radius is set by dividing the number of variants in common (from the output of the coordination step) by 12000. Note that for the coordination step we provided the median sample size as input value for *N*. For the LDpred step we applied the following thresholds for fraction of variants with non-zero effect (in addition to the default infinitesimal model): --PS=0.01,0.05,0.1,0.2,0.5. We used the plink2 software package for generating the PGSs by applying the --score option to the input weighted effect sizes and the genotype data set. As scoring genotype data sets, we used the entire NTR sample. PGSs were generated for all the NTR participants who had genotype data available.

### Between-family analysis

A between-family design was applied to test whether the outcome measures can be predicted by the PGSs for Morningness and RA. Given the exploratory nature of the associations, the results for all the threshold values are reported (*p* = 0.01, *p* = 0.05, *p* = 0.1, *p* = 0.2 and *p* = 0.3, *p* = 0.5 and *p* = infinity). We performed Generalized Estimation Equation modeling (GEE, Halekoh et al., 2006) with a conditional covariance matrix to account for the fact that observations for family members are dependent. The following covariates were included in the analyses: sex, age, age^2^, the first 10 principal components, and the survey wave. We handled a conservative Bonferroni-corrected significance value (0.05/126 = 0.0004).

If the PGSs for Morningness and RA were able to significantly predict the DSM *Depressive Symptoms* sum scores, we would perform a sensitivity analysis. Here we regressed the fourteen individual depressive symptoms items (see Table S1) on the PGSs for Morningness and RA. This time, we only included the PGSs with p-value thresholds that were significant in the prior GEE. The sensitivity analysis tested two hypotheses. Hypothesis 1: the PGSs predict all the depressive symptoms, possibly through a common latent factor (‘depression’). Hypothesis 2: the PGSs only predict the depressive symptoms that are directly related to sleep (symptoms 54, 77 & 100). Both hypotheses are visually represented in Figure 1.

**Figure 1.**
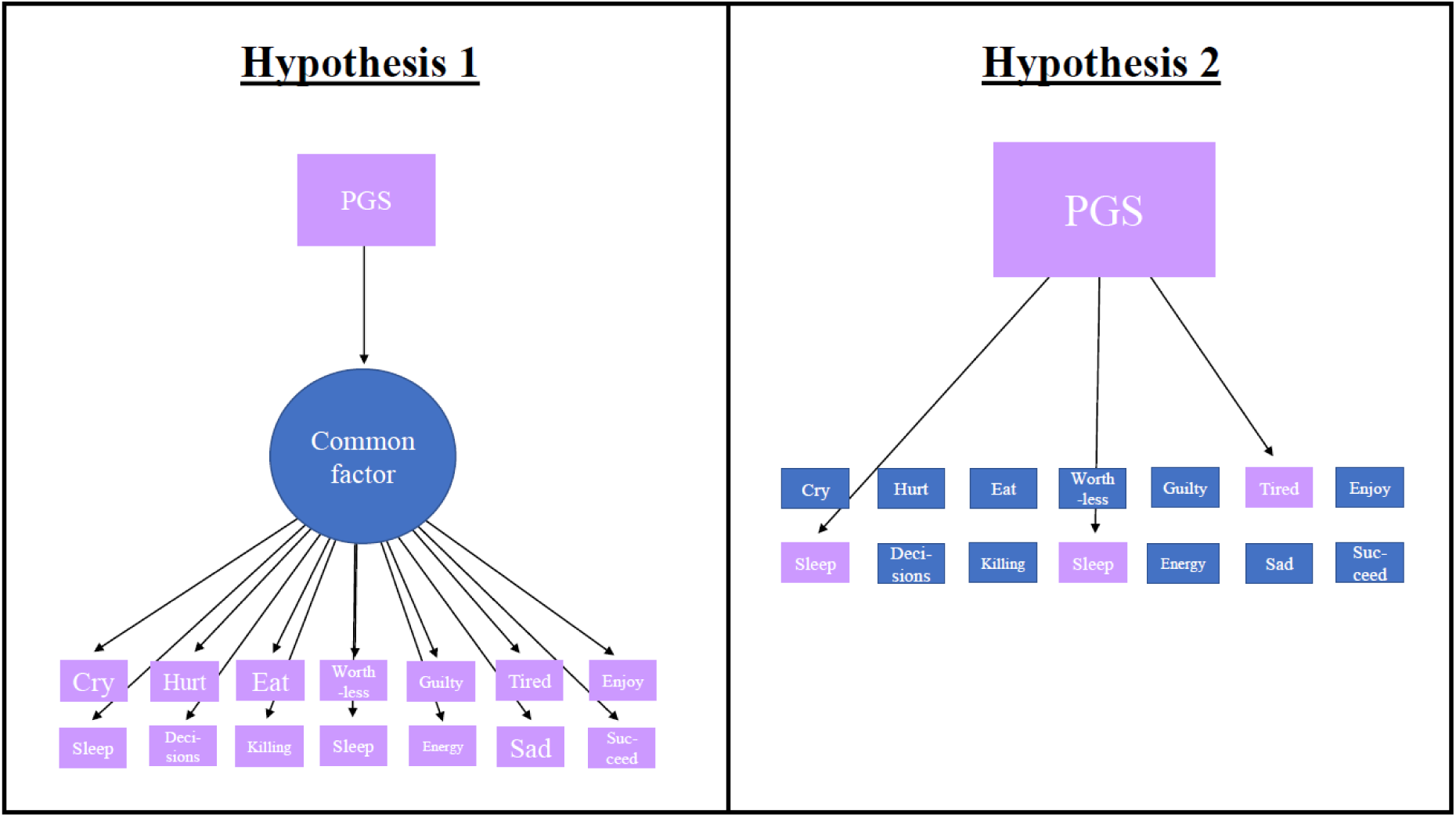
Hypotheses for the sensitivity analysis of Depressive Symptoms. Hypothesis 1: the PGSs predict all the separate depressive symptoms, possibly through a common latent factor. Hypothesis 2: the PGSs only predict the depressive symptoms that are directly related to sleep.

### Within-family analysis

As a follow-up analysis, we applied a within-family genetic design to detect passive gene-environment correlations (prGE). The theory behind this design is that parents generate an environment for their offspring that is correlated to their own genotypes (passive gene-environmental correlation). That is why an estimate of the effect of a PGS on an outcome not otherwise adjusted for the family environment is likely to include both direct genetic effects and indirect effects through the environment parents created. We can disentangle these direct genetic and indirect effects by using a within-family design applied to a subset containing only dizygotic (DZ) twin pairs. The related formula is shown below:

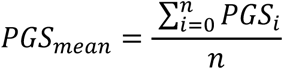

And the personal deviation for sibling i from the family mean as:

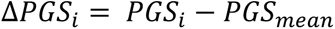

Then we can regress the outcome y on *X_mean_* and Δ_*i*_ as:

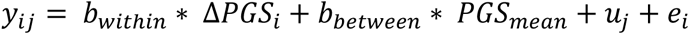

The intuitive reasoning behind the model is as follows: 1) siblings share the same parental genetic influences that have shaped their environment, 2) using within-family further rules out bias due to population stratification and assortative mating (Selzam et al., 2019) and 3) using DZ twins rather than sibling further controls for age, parental age, family income at a given age, and age-specific neighbourhood factors (Selzam et al., 2019). Based on this we expect the beta for the within-family predictions to be smaller than the beta for the between-family predictions.

We only applied the within-family analysis to the predictions that were found significant in the between-family analysis. Ninety-five percent confidence intervals (CI’s) were estimated using 1000 bootstrap resampling with replacement.

## Results

### Between-family analysis

Descriptive statistics are presented in Table S2. The results for the between-family analysis are summarized in Table 1. The PGS for Morningness significantly predicted two outcome measures: Morningness (*R*^2^ = 1.55%, *p* = < 0.001) and Depressive Symptoms (*R*^2^= 0.22%, *p* = < 0.001, Figure 2). This means that having a higher genetic predisposition for Morningness is related to being a morning person, and to experiencing fewer depressive symptoms. The *p*-value threshold of 0.2 had the best predictive value. The results for all the PGS Morningness analyses can be found in Table S3.

**Figure 2.**
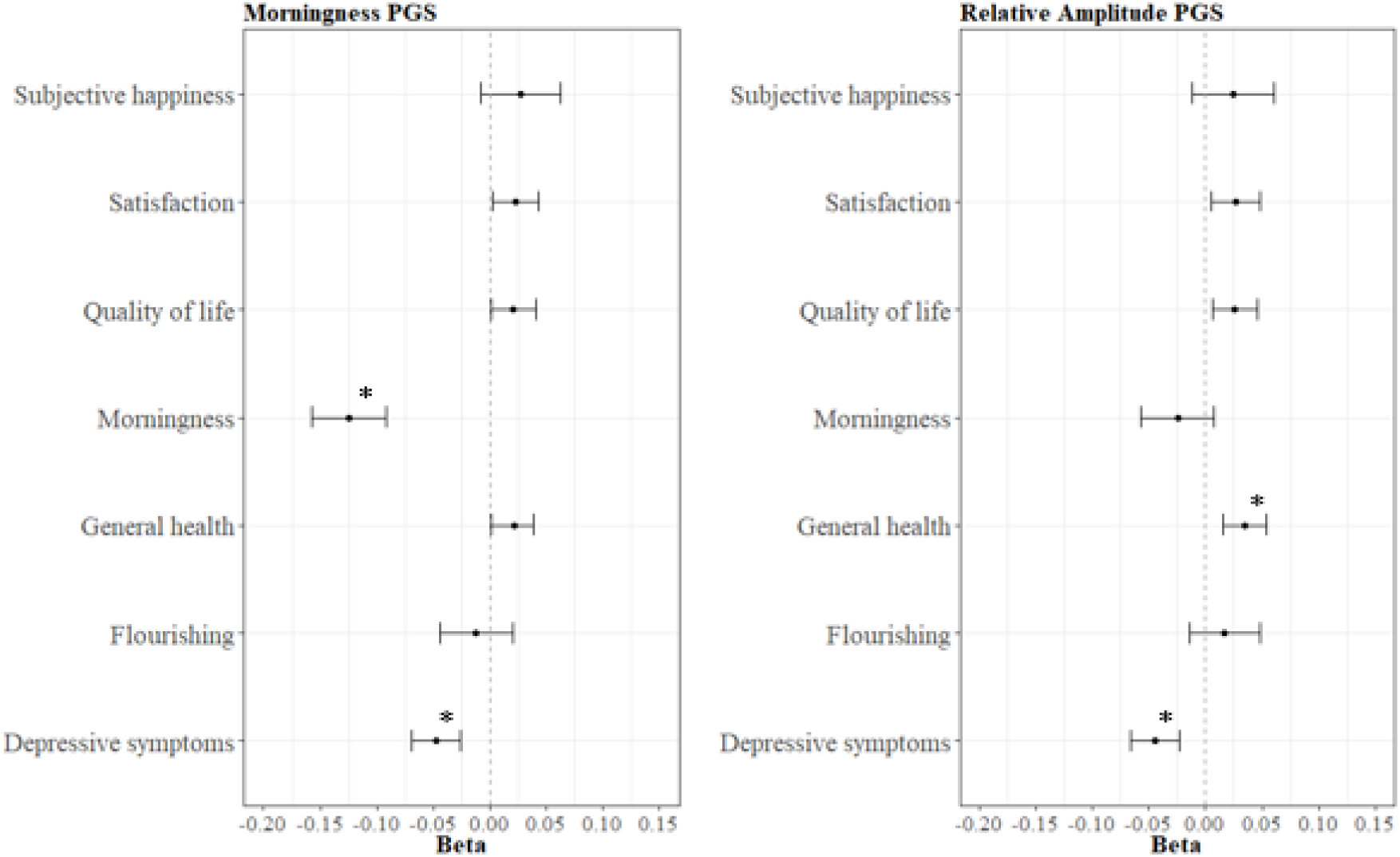
The seven outcome measures regressed on the PGS for Morningness and the PGS for Relative Amplitude. The * indicates a significant association.

**Table 1.** Summarized results from the between-family predictions for both the Morningness PGS and the RA PGS. This summary table shows the results for the analyses with the p-value thresholds that yielded the most explained variance (R^2^).

| **PGS** | **Phenotype** | ***b*** | ***SE*** | ***95% CI*** | **$R^2$** | ***p*** |
| --- | --- | --- | --- | --- | --- | --- |
| PGS for Morningness<br>($p$-value threshold) | | | | | | |
| 0.01 | Satisfaction with Life<br>($N = 9922$) | 0.0225 | 0.01 | 0.00 – 0.04 | 0.0005 | 0.0317 |
| 0.01 | Subjective Happiness<br>($N = 3222$) | 0.0276 | 0.02 | -0.01 – 0.06 | 0.0008 | 0.1242 |
| Infinity | Quality of Life<br>($N = 10431$) | 0.0224 | 0.01 | 0.00 – 0.04 | 0.0005 | 0.0268 |
| Infinity | Flourishing<br>($N = 4150$) | -0.0141 | 0.02 | -0.05 – 0.02 | 0.0002 | 0.3720 |
| Infinity | Health<br>($N = 10912$) | 0.0270 | 0.01 | 0.01 – 0.05 | 0.0007 | 0.0065 |
| 0.2 | Morningness<br>($N = 4048$) | -0.1246 | 0.02 | -0.16 – -0.09 | 0.0155 | 0.0000 |
| Infinity | ASR<br>($N = 9449$) | -0.0492 | 0.01 | -0.07 – -0.03 | 0.0024 | 0.0000 |
| PGS for RA<br>($p$-value threshold) | | | | | | |
| 0.01 | Satisfaction with Life<br>($N = 9922$) | 0.0266 | 0.01 | 0.01 – 0.05 | 0.0007 | 0.0135 |
| Infinity | Subjective Happiness<br>($N = 3222$) | 0.0254 | 0.02 | -0.01 – 0.06 | 0.0006 | 0.1619 |
| 0.01 | Quality of Life<br>($N = 10431$) | 0.0258 | 0.01 | 0.01 – 0.05 | 0.0007 | 0.0116 |
| Infinity | Flourishing<br>($N = 4150$) | 0.0198 | 0.02 | -0.01 – 0.05 | 0.0004 | 0.2202 |
| 0.01 | Health<br>($N = 10912$) | 0.0350 | 0.01 | 0.02 – 0.05 | 0.0012 | 0.0003 |
| 0.05 | Morningness<br>($N = 4048$) | -0.0247 | 0.02 | -0.06 – 0.01 | 0.0006 | 0.1366 |
| 0.01 | ASR<br>($N = 9449$) | -0.0447 | 0.01 | -0.07 – -0.02 | 0.0020 | 0.0000 |

**Note.** ASR = Depressive Symptoms sum score (Achenbach & Rescorla, 2003).

The PGS for RA also yielded two significant associations: Health (*R*^2^ = 0.12%, *p* = < 0.001) and Depressive Symptoms (*R*^2^ = 0.20%, *p* < 0.001, Figure 2). This means that having a genetic predisposition for a higher RA, meaning a well-functioning rest-activity rhythm, is associated with better health and less depressive symptoms. The *p*-value threshold of 0.01 had the best predictive value. The results for all the PGS RA analyses can be found Table S4.

### Sensitivity analysis: ASR items

Since both PGSs significantly predicted the DSM depressive symptoms sum scores, the 14 items were also separately regressed on the PGSs. Under the Bonferroni-corrected alpha, four of the predictions were significant. The top 3 predicted items for the PGS for Morningness were 1) *I have trouble making decisions* (*R*^2^ = 0.18, *p* = 0.0001), 2) *I do not have much energy* (*R*^2^ = 0.17, *p* = 0.0002), and 3) *I feel worthless or inferior* (*R*^2^ = 0.16, *p* = 0.0003). Higher PGS scores for Morningness predicted lower scores on these three items. The top 3 predicted items for the PGS for RA were 1) *I do not have much energy* (*R*^2^ = 0.21, *p* < 0.0001), *I think about killing myself* (*R*^2^ = 0.16, *ns*), 3), *I feel that I cannot succeed* (*R*^2^ = 0.15, *ns*). Higher PGS scores for RA predicted lower scores on these three items. The predictions are shown in Figure 3 and the detailed results for all the item analyses can be found in Table S5 & S6. The results contradict both hypotheses. The PGSs do not significantly predict all symptoms (hypothesis 1), nor do they only predict the sleep related symptoms (hypothesis 2). The results compared to the old hypotheses are depicted in Figure 4.

**Figure 3.**
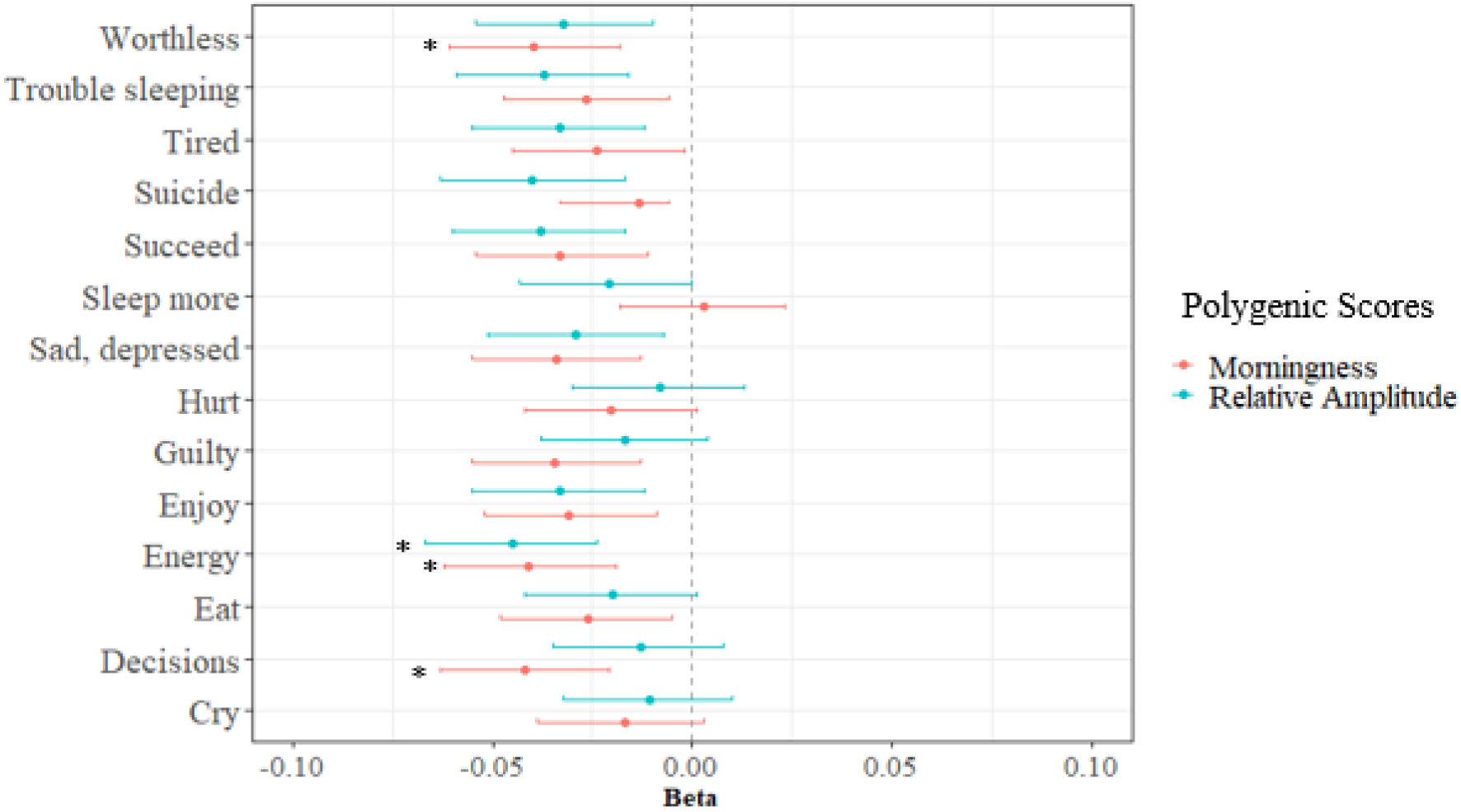
The fourteen Depressive Symptoms regressed on the PGS for Morningness and the PGS for Relative Amplitude. The * indicates a significant association.

**Figure 4.**
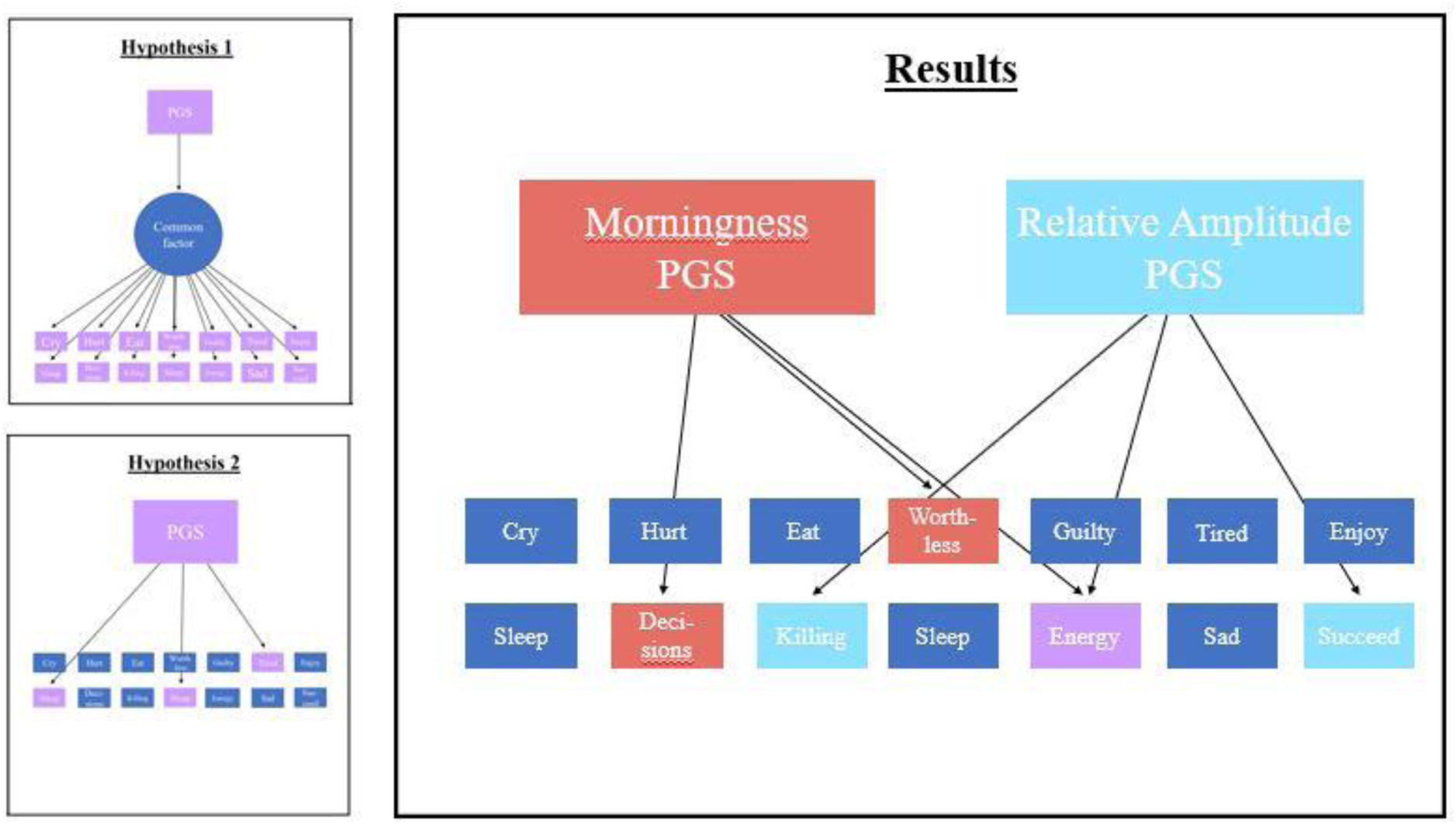
Hypotheses 1 and 2 compared to the results from the sensitivity analysis of Depressive Symptoms. The figure shows the top 3 predicted symptoms per PGS.

### Within-family analysis

Descriptive statistics for the within-family analysis can be found in Table S7. The analysis was bootstrapped with *N* = 1000. The within-family PGS predictions were only analyzed for the PGS associations that were statistically significant in the between-family PGS analysis. Therefore, Figures 5 depicts the between- and within-family analyses for 1) Morningness and the Depressive Symptoms sum score regressed on the PGS for Morningness, and 2) Self-Rated Health and the Depressive Symptoms sum score regressed on the PGS for RA. In the within-family analysis, the PGS for Morningness predicted Morningness (*b* = −0.18, CI (95%) = -.32, -.05). This means that twins with a higher PGS for Morningness are more likely to be a morning person than their co-twin. Because the regression coefficient was significantly different from zero, this suggests that there is gene-environment correlation. However, the CI’s of the within-family analysis overlap with the CI’s of the between-family analysis. The other within-family predictions did not yield significant results (Table S8 & S9).

**Figure 5.**
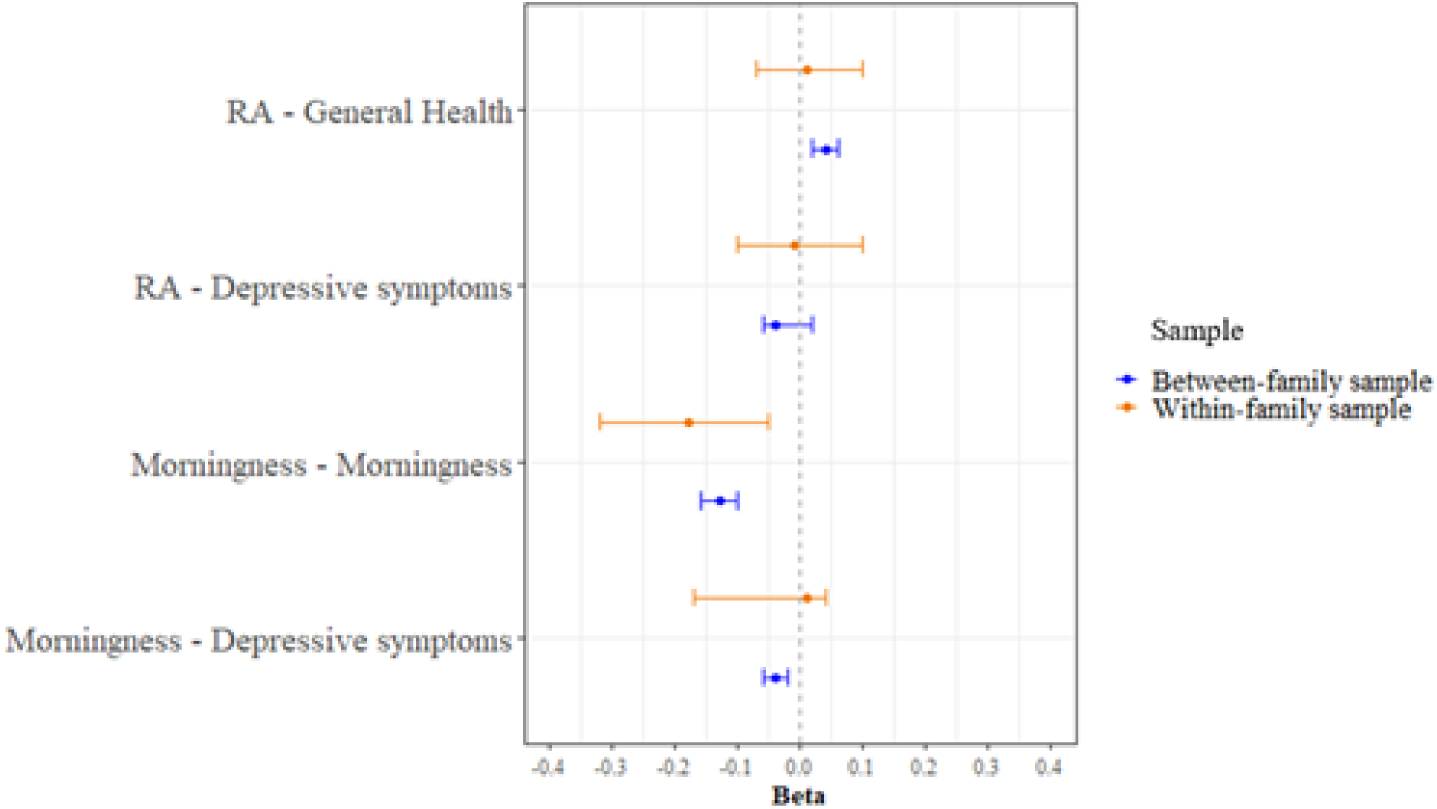
Bootstrap between- and within-family analyses results with 95% confidence intervals.

## Discussion

It is established that inappropriate and disruptive light exposure at night or too little time during the day, affects people’s circadian rhythm, health, and wellbeing (Lazzerini Ospri et al., 2017; LeGates et al., 2014). The goal of the study was to see to what extend the genetic predisposition for circadian rhythm related phenotypess like Morningness and Relative Amplitude reflecting rest-activity rhythm, captured with a PGS, could be used to predict wellbeing, depressive symptoms, chronotype, and health. We show that people with a higher genetic predisposition for being a morning person are more likely to be morning people and to have fewer depressive symptoms. People with a higher genetic predisposition for a high Relative Amplitude, reflecting a healthier rest-activity rhythm, are more likely to have good general health and also to have fewer depressive symptoms. We found no associations between the PGSs and wellbeing. Using sensitivity analyses it was tested if the PGSs predict all the depressive symptoms, possibly through a common latent factor (‘depression’) (Hypothesis 1) or if the PGSs only predict the depressive symptoms that are directly related to sleep (symptoms 54, 77 & 100, see Table S1; Hypothesis 2). The results did not confirm either hypothesis. The results from the within-family analysis suggest the presence of gene-environment correlation effects.

The result that people with a higher genetic predisposition for being a morning person and with a genetic predisposition for a high Relative Amplitude score were likely to have fewer depressive symptoms is in line with the results from Fergusson et al., (2018) where the PGSs for low Relative Amplitude predicted major depressive disorder, and the results from Jones et al., (2019) where they found that Morningness and Depressive Symptoms are partially influenced by the same genes, reflected in a significant genetic correlation of −0.16. On the contrary, the lack of associations of the PRS with wellbeing measures was unexpected. Jones at al., (2019) showed through both genetic correlation analysis and mendelian randomization analysis that Morningness and subjective wellbeing are related to each other. One explanation for this could be that the strict Bonferroni correction reduced power to detect a true positive effect. Considering the sign of the effects and disregarding the significance of the effects for a moment, our results show a pattern of a positive association between the PGSs and the wellbeing measures. In general, this association was stronger for the PGS for Relative Amplitude than for the PGS for Morningness.

The PGSs do not, in contrast to our hypotheses, relate to all depressive symptoms, nor do they only relate to the sleep related symptoms. These results are inconsistent with an influence of these PGS on depression symptoms via a common (latent) depression factor. Our findings are potentially complementary to the *causal systems perspective* (Borsboom, 2008). This perspective argues that the co-occurrence of symptoms is not driven by a common factor, but that this co-occurrence exists because of causal meaningful symptom-to-symptom interactions. Based on this absence of a common causal factor, it is likely that each symptom has their own psychological, neurological, and genetic root (Borsboom, 2008). Particularly the latter relates to our results. For example, the PGS for Morningness seems to relate more strongly to ‘*I have trouble making decisions’* than to *‘I cry a lot’*. At the same time ‘*I think of killing myself’* seems to be influenced by the genetic variants related to Relative Amplitude but not at all by the genetic variants related to Morningness. These are two examples that demonstrate differences in genetic root in depressive symptoms. This also suggests that although the two types of PGS both are derivatives of the construct of circadian rhythm, they may appeal very differently to these symptoms. Possible explanations for this could be that Morningness and Relative Amplitude are very distinct aspects of the circadian rhythm, or that Morningness is a subjective measure while Relative Amplitude is an objective measure. However, it is important to remember that we tested whether the PGS predictions were different from zero, but we did not formally test whether the PGS predictions were significantly different from each other. Our conclusions are therefore merely suggestive, or even merely an illustration of how, with increased power, future studies could explore the relation between circadian rhythms and depression under different models of depression.

The within-family analysis yielded unexpected results. There was a relation between the PGS for Morningness and the trait Morningnness. This indicates that, while correcting for effects of the family environment, the differences in outcome measures are related to polygenic differences between twins. This implies that differences in Morningness between dizygotic twins of the same family are driven by genetic differences between them. When comparing these within-family results to the (bootstrapped) between-family results, we expected that the within-family analysis would result in a smaller beta. This would then imply a gene-environment correlation (Selzam et al., 2019). The results showed the opposite: the beta for the within-family analysis was bigger compared to the between-family analysis. This could mean that the association between one’s genetic predisposition of being a morning person and actually being a morning person becomes stronger when we remove the effect of the family environment. However, even though the within-family prediction was significantly different from zero, it was not statistically different from the between-family prediction. This calls for caution when interpreting the result as a gene-environment correlation. This lack of statistical difference is likely to be an issue of power. For example, Okbay et al., (2022) compared within- and between-family PGS results for education attainment and found that about half of the predictive value of the PGS could be attributed to direct genetic effects from the within-family design. This also means that the other half could be attributed to assortative mating and population stratification and therefore their results justify gene-environment correlation. Compared to this study, Okbay et al., (2022) used a much larger sample (*n* = 56.500 vs 2100 for the within-family analysis) so it would be interesting to see how our findings would hold in a larger sample.

Zooming in on the within-family Morningness association, studies by Yamazaki (2007) and Leonhard & Randler (2009) have reflected on this role of the family environment on the circadian rhythm. Yamazaki (2007) showed that the circadian rhythm of first-time mothers is influenced by their few-months old children. Leonhard & Randler (2009) expended this and investigated how the circadian rhythm of women is influenced by both their children and their partners. Their results showed that the circadian rhythm and the chronotype of mothers is significantly influenced by their children, also when the children are on average 3.3 years old. Additionally, the synchrony between the mother and the child is stronger than between the mother and the partner, which indicates that children are a more influential social synchronizing factor than partners. Based on their data they argue that it is likely that the mother wakes up early because of the child, but that this does not necessarily reflect her own chronotype. Chronotypes of partners were moderately correlated (Leonhard & Randler., 2009), leaving the question whether this is due to assortative mating or the effect of living together. More recently, Pereira-Morales et al., (2019) published results on the association between individual chronotypes and perceived family chronotypes. Their findings show that the perceived family chronotype explains part of the variance in individual chronotype. Our result from the within-family analysis in Morningness leans towards these other findings. A follow-up study could include a family design with both parents and children to look at 1) the phenotypic associations of circadian rhythm and the direction of the association and at 2) the associations between the children’s PRS, the parental PRS (transmitted and non-transmitted) and the phenotypic circadian rhythm outcomes. The value of studying both the transmitted PRS and non-transmitted PRS is that they both reflect different types of effect. The transmitted parental PRS consists of alleles that are directly transmitted to the child, and therefore reflect a direct genetic effect. The non-transmitted parental PGS consists of alleles that were not transmitted to the child, but still have an effect on their children through the family environment. The latter is also known as the ‘genetic nurture’ phenomenon. These analyses together will allow us to better understand the role of genetic nature and family effects in circadian rhythm.

The present study was subjected to a number of limitations related to both the PGSs and the phenotypic data. One limitation regarding the PGSs is the lack of power in the PGS. Jones at al., found 351 SNP hits in a sample of 449,734 participants and Ferguson et al., (2018) found 5 SNP hits in a sample of 71,500 participants. GWASs of Morningness and Relative Amplitude with larger sample sizes are likely to find more SNPs and will therefore increase the predictive power of the PGSs. The relatively low power could be an explanation for the lack of association with wellbeing in this study. Besides limitations to the PGSs, there are also limitations to the phenotypic data. We identified two possible confounders in our data. The first is the time of day of filling in the questionnaire (morning/evening) and whether this is in line with the participant’s chronotype. It is likely that one will obtain different wellbeing scores from a ‘night-owl’-participant in the early morning compared to the evening. The second is the possible effect of seasonality. Some participants may be affected by the changing of the seasons (Patten et al., 2017; Wirz-Justice et al., 2019) and this may be reflected in e.g. their wellbeing data. To conclude, our study is limited by two other phenomena that are common in biological research. First is the lack of informativity of the PGSs. In addition to the predictive value being low, the PGSs do not provide us with information regarding the direct or indirect (causal) pathways between the relevant genes and the outcome measures (Plomin & von Stumm, 2022). Second, our study includes data from a sample with a predominantly European ancestry and with a high social-economic status. Therefore our results are not generalizable to other populations.

This study has several implications considering the relationship between circadian rhythm and mental health. The first is that the genes that are found to be related to circadian rhythm (until today) predict depressive symptoms and not wellbeing outcomes. Research on derpression should therefore more often include circadian rhythm as a relevant factor when studying causes and prevention in depression, while the strong focus on the general positive effects of sleep on wellbeing should be studied in more detail. Second, this study has shown that it is insightful to investigate different derivatives of circadian rhythm, since these have been shown to have a different genetic make-up and are likely to be associated with different traits. Therefore, different derivatives should be considered when trying to understand a certain trait. Third, even though this study did not directly proof gene-environment correlations in the circadian rhythm, the results for Morningness in the within-family design are promising enough to further investigate family effects in the circadian rhythm using genetically informed designs. And finally, the sensitivity analysis results advocate including item-based analysis when studying complex traits. Given that each symptom is likely to have a different genetic make-up and may be subjected to symptom-symptom interactions, item-based analyses are likely to provide researchers with new insights.

To conclude, this study showed that people with a genetic predisposition of being a morning person or a high relative amplitude are likely to have fewer depressive symptoms. The four strongest associated depressive symptoms described symptoms related to decision making, energy, and feeling worthless, rather than sleep. Our findings plead for a substantial role for the circadian rhythm in depression research, and to further explore the gene-environment correlation in circadian rhythm measures.

## Data Availability

All data produced in the present study are available upon reasonable request to the authors.

## Notes

### Competing Interest Statement

The authors have declared no competing interest.

### Funding Statement

This study was funded by the European Union's Horizon 2020 research and innovation programme under grant agreement no. 945238, and by an European Research Council (ERC) consolidator grant (WELL-BEING 771057 PI Bartels)

### Author Declarations

The Central Ethics Committee Research Involving Human Subjects of the VU University Medical Centre Amsterdam, an Institutional Review Board certified by the U.S. Office of Human Research Protections (IRB number IRB-2991 under Federal-wide Assurance-3703; IRB/institute codes, NTR 03-180) gave ethical approval for this work.

