## Supplementary material for "Using Polygenic Scores for Circadian Rhythm to predict Wellbeing, Depressive Symptoms, Chronotype, and Health": Table S1

### Supplementary materials

Table S1. *The fourteen items of the ASEBA-ASR* DSM-oriented *depressive problems scale.*

| **Item number** | **Item description** | **Item number** | **Item description** |
|---|---|---|---|
| **14** | I cry a lot | **77** | I sleep more than most other people |
| **18** | I deliberately try to hurt or kill myself | **78** | I have trouble making decisions |
| **24** | I do not eat as well as I should | **91** | I think about killing myself |
| **35** | I feel worthless or inferior | **100** | I have trouble sleeping |
| **52** | I feel very guilty | **102** | I do not have much energy |
| **54** | I feel tired without good reason | **103** | I am unhappy, sad, or depressed |
| **60** | There is very little that I enjoy | **107** | I feel that I cannot succeed |

**Table S2.**
*Descriptive statistics of all participants*

| **Variable** | **N** | ***M*** | ***SD*** |
|---|---|---|---|
| Sex | 12827 | 63.3% female | NA |
| Age | 12828 | 43.01 | 17.63 |
| Satisfaction with Life Scale | 9922 | 27.03 | 5.32 |
| Subjective Happiness Scale | 3222 | 22.66 | 4.50 |
| Quality of Life Scale | 10431 | 7.73 | 1.10 |
| Short Flourishing Scale | 4150 | 46.29 | 6.06 |
| Self-rated Health | 10912 | 3.97 | 0.75 |
| Morningness | 4048 | 3.15 | 1.28 |

| | | | |
|---|---|---|---|
| Depressive Problems Scale | 9449 | 3.77 | 3.77 |
| ASR item 14: “ I cry a lot” | 9013 | 0.20 | 0.44 |
| ASR item 18: “ I deliberately try to hurt or kill myself” | 8861 | 0.02 | 0.14 |
| ASR item 24:“ I do not eat as well as I should” | 8839 | 0.45 | 0.60 |
| ASR item 35:“ I feel worthless or inferior” | 8992 | 0.21 | 0.46 |
| ASR item 52:“ I feel very guilty” | 8993 | 0.19 | 0.43 |
| ASR item 54:“ I feel tired without good reason” | 8816 | 0.47 | 0.62 |
| ASR item 60:“ There is very little that I enjoy” | 8813 | 0.13 | 0.38 |
| ASR item 77:“ I sleep more than most other people” | 8811 | 0.18 | 0.48 |
| ASR item 78:“ I have trouble making decisions” | 8820 | 0.48 | 0.59 |
| ASR item 91:“ I think about killing myself” | 8974 | 0.03 | 0.20 |
| ASR item 100:“ I have trouble sleeping” | 8779 | 0.36 | 0.62 |
| ASR item 102:“ I do not have much energy” | 8785 | 0.46 | 0.62 |
| ASR item 103:“ I am unhappy, sad, or depressed” | 8949 | 0.23 | 0.47 |
| ASR item 107:“ I feel that I cannot succeed” | 8948 | 0.31 | 0.52 |

**Table S3.**

*Phenotypes regressed on the PGS for Morningness*

| PGS for Morningness | *b* | *SE* | *95% CI* | $R^2$ | *p* |
|---|---|---|---|---|---|
| **Satisfaction with Life (N = 9922)** | | | | | |
| 0.01 | 0.0225 | 0.01 | 0.00 – 0.04 | 0.0005 | 0.0317 |
| 0.05 | 0.0164 | 0.01 | -0.00 - 0.04 | 0.0003 | 0.1180 |
| 0.1 | 0.0176 | 0.01 | -0.00 – 0.04 | 0.0003 | 0.0933 |
| 0.2 | 0.0170 | 0.01 | -0.00 – 0.04 | 0.0003 | 0.1065 |
| 0.3 | 0.0170 | 0.01 | -0.00 – 0.04 | 0.0003 | 0.1047 |
| 0.5 | 0.0168 | 0.01 | -0.00 – 0.04 | 0.0003 | 0.1111 |
| Infinity | 0.0186 | 0.01 | -0.00 – 0.04 | 0.0003 | 0.0791 |
| **Subjective Happiness (N = 3222)** | | | | | |
| 0.01 | 0.0276 | 0.02 | -0.01 – 0.06 | 0.0008 | 0.1242 |
| 0.05 | 0.0061 | 0.02 | -0.03 – 0.04 | 0.0000 | 0.7261 |
| 0.1 | 0.0086 | 0.02 | -0.03 – 0.04 | 0.0001 | 0.6302 |
| 0.2 | 0.0104 | 0.02 | -0.02 – 0.05 | 0.0001 | 0.5587 |
| 0.3 | 0.0112 | 0.02 | -0.02 – 0.05 | 0.0001 | 0.5294 |
| 0.5 | 0.0120 | 0.02 | -0.02 – 0.05 | 0.0001 | 0.7399 |
| Infinity | 0.0104 | 0.02 | -0.03 – 0.05 | 0.0001 | 0.5659 |
| **Quality of Life (N = 10431)** | | | | | |
| 0.01 | 0.0193 | 0.00 | 0.00 – 0.04 | 0.0004 | 0.0525 |
| 0.05 | 0.0152 | 0.01 | -0.00 – 0.04 | 0.0002 | 0.1323 |
| 0.1 | 0.0203 | 0.01 | 0.00 – 0.04 | 0.0004 | 0.0453 |
| 0.2 | 0.0208 | 0.01 | 0.00 – 0.04 | 0.0004 | 0.0403 |
| 0.3 | 0.0203 | 0.01 | 0.00 – 0.04 | 0.0004 | 0.0455 |
| 0.5 | 0.0205 | 0.01 | 0.00 – 0.04 | 0.0004 | 0.0438 |
| Infinity | 0.0224 | 0.01 | 0.00 – 0.04 | 0.0005 | 0.0268 |
| **Flourishing (N = 4150)** | | | | | |
| 0.01 | 0.0064 | 0.02 | -0.03 – 0.04 | 0.0000 | 0.6909 |
| 0.05 | -0.0112 | 0.02 | -0.04 – 0.02 | 0.0001 | 0.4736 |

| | | | | | |
|---|---|---|---|---|---|
| 0.1 | -0.0112 | 0.02 | -0.04 – 0.02 | 0.0001 | 0.4788 |
| 0.2 | -0.0129 | 0.02 | -0.04 – 0.02 | 0.0002 | 0.4110 |
| 0.3 | -0.0123 | 0.02 | -0.04 – 0.02 | 0.0002 | 0.4317 |
| 0.5 | -0.0122 | 0.02 | -0.04 – 0.02 | 0.0001 | 0.4382 |
| Infinity | -0.0141 | 0.02 | -0.05 – 0.02 | 0.0002 | 0.3720 |
| **Self-Rated Health (N = 10912)** | | | | | |
| 0.01 | 0.0208 | 0.01 | 0.00 – 0.04 | 0.0004 | 0.0274 |
| 0.05 | 0.0220 | 0.01 | 0.00 – 0.04 | 0.0004 | 0.0397 |
| 0.1 | 0.0187 | 0.01 | -0.00 – 0.04 | 0.0003 | 0.0546 |
| 0.2 | 0.0203 | 0.01 | 0.00 – 0.04 | 0.0004 | 0.0384 |
| 0.3 | 0.0199 | 0.01 | 0.00 – 0.04 | 0.0004 | 0.0432 |
| 0.5 | 0.0203 | 0.01 | 0.00 – 0.04 | 0.0004 | 0.0389 |
| Infinity | 0.0270 | 0.01 | 0.01 – 0.05 | 0.0007 | 0.0065 |
| **Morningness (N = 4048)** | | | | | |
| 0.01 | -0.0326 | 0.02 | -0.07 – -0.00 | 0.0011 | 0.0484 |
| 0.05 | -0.0829 | 0.02 | -0.11 – 0.05 | 0.0069 | 0.0000 |
| 0.1 | -0.1201 | 0.02 | -0.15 – -0.09 | 0.0144 | 0.0000 |
| 0.2 | -0.1246 | 0.02 | -0.16 – -0.09 | 0.0155 | 0.0000 |
| 0.3 | -0.1232 | 0.02 | -0.16 – -0.09 | 0.0152 | 0.0000 |
| 0.5 | -0.1232 | 0.02 | -0.16 – -0.09 | 0.0152 | 0.0000 |
| Infinity | -0.1139 | 0.02 | -0.15– 0.08 | 0.0130 | 0.0000 |
| **DSM-oriented *depressive problems scale* (N = 9449)** | | | | | |
| 0.01 | -0.0336 | 0.01 | -0.05 – -0.01 | 0.0011 | 0.0019 |
| 0.05 | -0.0303 | 0.01 | -0.05 – -0.01 | 0.0009 | 0.0043 |
| 0.1 | -0.0461 | 0.01 | -0.07 – -0.02 | 0.0021 | 0.0000 |
| 0.2 | -0.0473 | 0.01 | -0.07 – -0.03 | 0.0022 | 0.0000 |
| 0.3 | -0.0471 | 0.01 | -0.07 – -0.03 | 0.0022 | 0.0000 |
| 0.5 | -0.0462 | 0.01 | -0.07 – -0.03 | 0.0021 | 0.0000 |
| Infinity | -0.0492 | 0.01 | -0.07 – -0.03 | 0.0024 | 0.0000 |

**Table S4.**

*Phenotypes regressed on the PGS for RA*

| PGS for RA | *b* | *SE* | *95% CI* | $R^2$ | *p* |
|---|---|---|---|---|---|
| **Satisfaction with Life (N = 9922)** | | | | | |
| 0.01 | 0.0266 | 0.01 | 0.01 – 0.05 | 0.0007 | 0.0135 |
| 0.05 | 0.0249 | 0.01 | 0.00 – 0.05 | 0.0006 | 0.0210 |
| 0.1 | 0.0247 | 0.01 | 0.00 – 0.05 | 0.0006 | 0.0220 |
| 0.2 | 0.0246 | 0.01 | 0.00 – 0.05 | 0.0006 | 0.0225 |
| 0.3 | 0.0246 | 0.01 | 0.00 – 0.05 | 0.0006 | 0.0226 |
| 0.5 | 0.0246 | 0.01 | 0.00 – 0.05 | 0.0006 | 0.0228 |
| Infinity | 0.0246 | 0.01 | 0.00 – 0.04 | 0.0006 | 0.0221 |
| **Subjective Happiness (N = 3222)** | | | | | |
| 0.01 | 0.0246 | 0.02 | -0.01 – 0.06 | 0.0006 | 0.1797 |
| 0.05 | 0.0228 | 0.02 | -0.01 – 0.06 | 0.0005 | 0.2142 |
| 0.1 | 0.0226 | 0.02 | -0.01 – 0.06 | 0.0005 | 0.2185 |
| 0.2 | 0.0224 | 0.02 | -0.01 – 0.06 | 0.0005 | 0.2212 |
| 0.3 | 0.0224 | 0.02 | -0.01 – 0.06 | 0.0005 | 0.2217 |
| 0.5 | 0.0224 | 0.02 | -0.01 – 0.06 | 0.0005 | 0.2230 |
| Infinity | 0.0254 | 0.02 | -0.01 – 0.06 | 0.0006 | 0.1619 |
| **Quality of Life (N = 10431)** | | | | | |
| 0.01 | 0.0258 | 0.01 | 0.01 – 0.05 | 0.0007 | 0.0116 |
| 0.05 | 0.0246 | 0.01 | 0.00 – 0.04 | 0.0006 | 0.0161 |
| 0.1 | 0.0245 | 0.01 | 0.00 – 0.04 | 0.0006 | 0.0167 |
| 0.2 | 0.0244 | 0.01 | 0.00 – 0.04 | 0.0006 | 0.0170 |
| 0.3 | 0.0244 | 0.01 | 0.00 – 0.04 | 0.0006 | 0.0171 |
| 0.5 | 0.0244 | 0.01 | 0.00 – 0.04 | 0.0006 | 0.0172 |
| Infinity | 0.0181 | 0.01 | -0.00 – 0.04 | 0.0003 | 0.0836 |
| **Flourishing (N = 4150)** | | | | | |
| 0.01 | 0.0170 | 0.02 | -0.01 – 0.05 | 0.0003 | 0.2857 |
| 0.05 | 0.0156 | 0.02 | -0.02 – 0.05 | 0.0002 | 0.3269 |
| 0.1 | 0.0153 | 0.02 | -0.02 – 0.05 | 0.0002 | 0.3351 |
| 0.2 | 0.0152 | 0.02 | -0.02 – 0.05 | 0.0002 | 0.3387 |
| 0.3 | 0.0152 | 0.02 | -0.02 – 0.05 | 0.0002 | 0.3398 |
| 0.5 | 0.0152 | 0.02 | -0.02 – 0.05 | 0.0002 | 0.3404 |

| | | | | | |
|---|---|---|---|---|---|
| Infinity | 0.0198 | 0.02 | -0.01 – 0.05 | 0.0004 | 0.2202 |
| **Health (N = 10912)** | | | | | |
| 0.01 | 0.0350 | 0.01 | 0.02 – 0.05 | 0.0012 | 0.0003 |
| 0.05 | 0.0336 | 0.01 | 0.01 – 0.05 | 0.0011 | 0.0006 |
| 0.1 | 0.0335 | 0.01 | 0.01 – 0.05 | 0.0011 | 0.0006 |
| 0.2 | 0.0334 | 0.01 | 0.01 – 0.05 | 0.0011 | 0.0006 |
| 0.3 | 0.0334 | 0.01 | 0.01 – 0.05 | 0.0011 | 0.0006 |
| 0.5 | 0.0333 | 0.01 | 0.01 – 0.05 | 0.0011 | 0.0006 |
| Infinity | 0.0332 | 0.01 | 0.01 – 0.05 | 0.0011 | 0.0008 |
| **Morningness (N = 4048)** | | | | | |
| 0.01 | -0.0241 | 0.02 | -0.06 – 0.01 | 0.0006 | 0.1468 |
| 0.05 | -0.0246 | 0.02 | -0.06 – 0.01 | 0.0006 | 0.1377 |
| 0.1 | -0.0246 | 0.02 | -0.06 – 0.01 | 0.0006 | 0.1388 |
| 0.2 | -0.0246 | 0.02 | -0.06 – 0.01 | 0.0006 | 0.1388 |
| 0.3 | -0.0246 | 0.02 | -0.06 – 0.01 | 0.0006 | 0.1378 |
| 0.5 | -0.0247 | 0.02 | -0.06 – 0.01 | 0.0006 | 0.1366 |
| Infinity | -0.0246 | 0.02 | -0.06 – 0.01 | 0.0006 | 0.1307 |
| **ASR (N = 9449)** | | | | | |
| 0.01 | -0.0447 | 0.01 | -0.07 – -0.02 | 0.0020 | 0.0000 |
| 0.05 | -0.0443 | 0.01 | -0.07 – -0.02 | 0.0020 | 0.0000 |
| 0.1 | -0.0442 | 0.01 | -0.07 – -0.02 | 0.0020 | 0.0000 |
| 0.2 | -0.0442 | 0.01 | -0.07 – -0.02 | 0.0020 | 0.0000 |
| 0.3 | -0.0442 | 0.01 | -0.07 – -0.02 | 0.0020 | 0.0000 |
| 0.5 | -0.0442 | 0.01 | -0.07 – -0.02 | 0.0020 | 0.0000 |
| Infinity | -0.0427 | 0.01 | -0.06 – -0.02 | 0.0018 | 0.0001 |

**Table S5.**

*Associations between the PGS for Morningness and the separate Depressive symptoms*

| **PGS for Morningness** | ***b*** | ***SE*** | ***95% CI*** | $R^2$ | ***p*** |
|---|---|---|---|---|---|
| 14- I cry a lot (N = 9013) | -0.0180 | 0.01 | -0.04 – 0.00 | 0.0003 | 0.0939 |
| 18 – I deliberately try to hurt or kill myself (N = 8861) | -0.0204 | 0.01 | -0.04 – 0.00 | 0.0004 | 0.0663 |
| 24 – I do not eat as well as I should (N = 8839) | -0.0267 | 0.01 | -0.05 – -0.01 | 0.0007 | 0.0157 |
| 35 – I feel worthless or inferior (N = 8992) | -0.0398 | 0.01 | -0.06 – -0.02 | 0.0016 | 0.0003 |
| 52 – I feel very guilty (N = 8993) | -0.0343 | 0.01 | -0.06 – -0.01 | 0.0012 | 0.0015 |
| 54 – I feel tired without good reason (N = 8816) | -0.0238 | 0.01 | -0.05 – -0.00 | 0.0006 | 0.0310 |
| 60 – There is very little that I enjoy (N = 8813) | -0.0310 | 0.01 | -0.05 – -0.01 | 0.0010 | 0.0055 |
| 77 – I sleep more than most other people (N = 8811) | 0.00263 | 0.01 | -0.02 – 0.02 | 0.0000 | 0.8038 |
| 78 – I have trouble making decisions (N = 8820) | -0.0421 | 0.01 | -0.06 – -0.02 | 0.0018 | 0.0001 |
| 91 – I think about killing myself (N = 8974) | -0.0134 | 0.01 | -0.03 – 0.01 | 0.0002 | 0.1854 |
| 100 – I have trouble sleeping (N = 8779) | -0.0267 | 0.01 | -0.05 – -0.01 | 0.0007 | 0.0126 |
| 102 -I do not have much energy (N = 8785) | -0.0410 | 0.01 | -0.06 – -0.02 | 0.0017 | 0.0002 |
| 103 – I am unhappy, sad or depressed (N = 8949) | -0.0340 | 0.01 | -0.06 – -0.01 | 0.0012 | 0.0017 |
| 107 – I feel that I cannot succeed (N = 8948) | -0.0327 | 0.01 | -0.05 – -0.01 | 0.0011 | 0.0031 |

**Table S6.**

*Associations between the PGS for RA and the separate Depressive symptoms*

| PRS for RA | *b* | *SE* | *95% CI* | $R^2$ | *p* |
|---|---|---|---|---|---|
| 14- I cry a lot (N = 9013) | -0.0171 | 0.01 | -0.03 – 0.01 | 0.0001 | 0.3175 |
| 18 – I deliberately try to hurt or kill myself (N = 8861) | -0.0083 | 0.01 | -0.03 – 0.01 | 0.0001 | 0.4547 |
| 24 – I do not eat as well as I should (N = 8839) | -0.0205 | 0.01 | -0.04 – .00 | 0.0004 | 0.0606 |
| 35 – I feel worthless or inferior (N = 8992) | -0.0321 | 0.01 | -0.05 – -0.01 | 0.0010 | 0.0037 |
| 52 – I feel very guilty (N = 8993) | -0.0172 | 0.01 | -0.04 – 0.00 | 0.0003 | 0.1075 |
| 54 – I feel tired without good reason (N = 8816) | -0.0333 | 0.01 | -0.05 – -0.01 | 0.0011 | 0.0024 |
| 60 – There is very little that I enjoy (N = 8813) | -0.0334 | 0.01 | -0.05 – -0.01 | 0.0011 | 0.0019 |
| 77 – I sleep more than most other people (N = 8811) | -0.0215 | 0.01 | -0.04 – -0.00 | 0.0005 | 0.0466 |
| 78 – I have trouble making decisions (N = 8820) | -0.0133 | 0.01 | -0.03 – 0.01 | 0.0002 | 0.2256 |
| 91 – I think about killing myself (N = 8974) | -0.0399 | 0.01 | -0.06 – -0.02 | 0.0016 | 0.0006 |
| 100 – I have trouble sleeping (N = 8779) | -0.0373 | 0.01 | -0.06 – -0.02 | 0.0014 | 0.0006 |
| 102 – I do not have much energy (N = 8785) | -0.0453 | 0.01 | -0.07 – -0.02 | 0.0021 | 0.0000 |
| 103 – I am unhappy, sad or depressed (N = 8949) | -0.0290 | 0.01 | -0.05 – -0.01 | 0.0008 | 0.0087 |
| 107 – I feel that I cannot succeed (N = 8948) | -0.0383 | 0.01 | -0.06 – -0.02 | 0.0015 | 0.0005 |

**Table S7.**

*Descriptive statistics of the DZ sample*

| **Variable** | ***N*** | ***M*** | ***SD*** |
|---|---|---|---|
| Sex | 2099 | 62.2% female | NA |
| Age | 2099 | 32.17 | 14.38 |
| Satisfaction with Life Scale | 1692 | 26.83 | 5.40 |
| Subjective Happiness Scale | 438 | 22.24 | 4.62 |
| Quality of Life Scale | 1681 | 7.60 | 1.13 |
| Short Flourishing Scale | 606 | 46.18 | 6.22 |
| Self-rated Health | 1746 | 4.03 | 0.73 |
| Morningness | 787 | 3.29 | 1.29 |
| Depressive Problems Scale | 1762 | 3.99 | 3.91 |
| ASR item 14: “ I cry a lot” | 1622 | 0.21 | 0.45 |
| ASR item 18: “ I deliberately try to hurt or kill myself” | 1598 | 0.02 | 0.17 |
| ASR item 24: “ I do not eat as well as I should” | 1594 | 0.51 | 0.62 |
| ASR item 35: “ I feel worthless or inferior” | 1616 | 0.24 | 0.49 |
| ASR item 52: “ I feel very guilty” | 1616 | 0.19 | 0.43 |
| ASR item 54: “ I feel tired without good reason” | 1582 | 0.50 | 0.64 |
| ASR item 60: “ There is very little that I enjoy” | 1587 | 0.14 | 0.39 |
| ASR item 77: “ I sleep more than most other people” | 1579 | 0.20 | 0.49 |
| ASR item 78: “ I have trouble making decisions” | 1584 | 0.53 | 0.62 |
| ASR item 91: “ I think about killing myself” | 1613 | 0.04 | 0.22 |
| ASR item 100: “ I have trouble sleeping” | 1576 | 0.34 | 0.59 |
| ASR item 102: “ I do not have much energy” | 1578 | 0.46 | 0.62 |
| ASR item 103: | 1607 | 0.24 | 0.49 |

| | | | |
|---|---|---|---|
| " I am unhappy, sad, or depressed" ASR item 107: " I feel that I cannot succeed" | 1608 | 0.33 | 0.55 |

**Table S8.**

*Bootstrap results for Morningness and Depressive Symptoms regressed on the PGS for Morningness in the between- and within-family sample.*

| **Morningness PRS (*p*-value threshold = 0.2)** | ***b (SE)*** | ***95% CI*** | ***Δ Beta*** | ***Δ SE*** | ***Δ CI*** |
|---|---|---|---|---|---|
| Morningness Between-family sample | -0.1275 (0.01) | -0.16 – -0.10 | 0.01 | 0.07 | -0.13 – 0.14 |
| Morningness Within-family sample | -0.1830 (0.07) | -0.32 – -0.05 | 0.06 | 0.07 | -0.08 – 0.19 |
| Depressive Symptoms Between-family sample | -0.0401 (0.01) | -0.06 – -0.02 | 0.01 | 0.05 | -0.09 – 0.10 |
| Depressive Symptoms Within-family sample | 0.01 (0.05) | -0.17 – 0.04 | 0.04 | 0.05 | -0.06 – 0.13 |

***Note***. Δ Beta = difference between the beta from the initial analysis and the beta in the bootstrap analysis. Δ SE = standard error between the differences between the between-family beta's and within-family beta's. Δ CI = confidence intervals between the Δ Beta based on the Δ SE.

**Table S9.**

*Bootstrap results for RA and Depressive symptoms regressed on the PGS for Morningness in the between- and within-family sample.*

| **RA PRS (*p*-value threshold = 0.01)** | ***b (SE)*** | ***95% CI*** | ***Δ Beta*** | ***Δ SE*** | ***Δ CI*** |
|---|---|---|---|---|---|
| Health Between-family sample | 0.0386 (0.01) | 0.02 – 0.06 | 0.01 | 0.05 | -0.09 – 0.10 |
| Health Within-family sample | 0.0144 (0.05) | -0.07 – 0.10 | 0.02 | 0.05 | -0.08 – 0.12 |
| Depressive Symptoms Between-family sample | -0.0440 (0.01) | -0.06 – 0.02 | 0.01 | 0.05 | -0.10 – 0.10 |
| Depressive Symptoms Within-family sample | -0.0038 (0.05) | -0.10 – 0.10 | 0.04 | 0.05 | -0.06 – 0.14 |

***Note***. Δ Beta = difference between the beta from the initial analysis and the beta in the bootstrap analysis. Δ SE = standard error between the differences between the between-family beta's and within-family beta's. Δ CI = confidence intervals between the Δ Beta based on the Δ SE.
